# DNM1-related disorder is characterized by recurrent variants and phenotypic homogeneity

**DOI:** 10.64898/2026.04.05.26350183

**Authors:** Alicia G. Harrison, Shiva Ganesan, Hongbo M. Xie, Shridhar Parthasarathy, Jillian L. McKee, Jan H. Magielski, Kim Thalwitzer, Rohit Lobo, Manuela Pendziwiat, Andreas van Baalen, Hiltrud Muhle, Annapurna Poduri, Alisa Mo, Gert Wiegand, Katrin Õunap, Ange-Line Bruel, Marcello Scala, Valeria Capra, Sarah M Ruggiero, Ingo Helbig

## Abstract

**Purpose:** *DNM1*-related disorder is a rare developmental and epileptic encephalopathy. The current understanding of the clinical spectrum is based on sparse patient descriptions. Here, we compile the largest *DNM1* cohort to date, to characterize the genotypic and phenotypic landscape of the disorder.

**Methods:** Phenotypic data was manually curated from 95 individuals from multiple sources and harmonized using the Human Phenotype Ontology framework.

**Results:** Disease-causing variants in *DNM1* cluster in mutational hotspots within the gene, which achieve ‘Strong’ and ‘Moderate’ evidence for pathogenicity based on ACMG guidelines. The overall *DNM1* phenotype was homogeneous compared to other genetic epilepsy conditions: *SCN2A, SCN8A, STXBP1*, and *SYNGAP1*. The p.R237W (n=15) variant was associated with bilateral tonic-clonic seizures, infantile spasms, and dystonia. The p.I398_R399insCR (n=14) variant was associated with severe hypotonia, profound global delay, and cortical visual impairment. Five individuals with homozygous loss-of-function variants were clinically similar to dominant-negative *DNM1*-related disorder, but microcephaly and brain MRI abnormalities were more common in this group.

**Conclusion:** A harmonized cohort of individuals with *DNM1*-related disorder was analyzed to define mutational hotspots and reveal novel genotype-phenotype correlations. Due to the homogeneous phenotype, disease mechanism, and high proportion of recurrent variants, *DNM1* represents an attractive target for targeted therapy development.

## Introduction

Gene discovery in the epilepsies has been a dynamic area of research that has transformed a group of previously obscure disorders into well-defined neurogenetic conditions with a diagnostic rate of up to 45%.^1^ The *DNM1* gene (HGNC:2972) is located on chromosome 9 and encodes the protein dynamin 1, a GTPase involved in synaptic vesicle fission on neuronal presynaptic membranes.^2^ Dynamin 1 self-assembles into tetrameric spirals around the neck of budding vesicles during receptor-mediated endocytosis.^3^ Initial functional studies demonstrated that pathogenic variants in *DNM1* resulted in depletion of synaptic vesicles.^4^

*De novo* variants in *DNM1* were first identified in patients with severe childhood epilepsy in large-scale genetic studies,^5^ and subsequently in clinical case reports.^6,7^ Von Spiczak et al. compiled the first *DNM1* cohort in 2017 and outlined the clinical phenotype of *DNM1*-related encephalopathy based on 21 individuals.^8^ The overall phenotype is known to include severe to profound intellectual disability, hypotonia, and an epilepsy which is characterized by infantile spasms that often evolve into Lennox-Gastaut Syndrome.^8^ Pathogenic missense variants reported in *DNM1* cluster in hotspots in the GTPase and Middle domains, which is consistent with the established dominant-negative mechanism of disease.^8^ Our initial publication characterizing *DNM1* found that one recurrent variant, p.R237W, accounted for 30% of the reported cases.^8^ Another highly recurrent variant, p.I398_R399insCR, was subsequently identified in exon 10a of *DNM1*, which creates an in-frame upstream splice acceptor that leads to the insertion of two amino acids, which is thought to impair oligomerization-dependent activity.^9^ The location of this variant is significant because exon 10a is included in the isoform most expressed in brain tissue but is excluded from the canonical transcript which has been largely used to study and report variants in *DNM1*.^9^

While most cases of *DNM1*-related disorder are caused by heterozygous missense variants, homozygous protein-truncating variants have been identified in multiple unrelated individuals with similar clinical features to those seen in individuals with pathogenic missense variants.^10-12^ This suggests that there are two molecular mechanisms of disease for *DNM1*-related disorder: heterozygous dominant-negative variants with an autosomal dominant pattern of inheritance and biallelic loss-of-function variants with an autosomal recessive pattern of inheritance.

Further characterization of the genotype-phenotype relationship in *DNM1*-related disorder is needed, and has been hindered, in part, by the small sample sizes included in most publications and the rarity of the condition. Here, we compile a large data set by leveraging the Human Phenotype Ontology to harmonize data drawn from multiple sources, an approach has been successful in several other conditions.^13,14^ Clear characterization of the phenotypic spectrum is important in providing anticipatory guidance to families. We also demonstrate that a computational approach can provide overarching insights about the relationships between genetic disorders. We find that *DNM1*-related disorder presents with a narrow and quantifiable clinical spectrum with recognizable clinical patterns that may allow for future precision medicine treatments.

## Methods

### Subject and phenotype extraction

A total of 95 individuals are included in this study. A review of the literature was performed to identify all reported individuals with *DNM1*-related disorder. Relevant publications were identified through the Human Gene Mutation Database (HGMD).^16^ 68 individuals were identified with available phenotype information. We also included 21 individuals recruited through the Epilepsy Genetics Research Project (EGRP) at Children’s Hospital of Philadelphia (CHOP) who had not been previously reported. An additional 6 previously unreported participants were identified through broad research protocols at University Medical Center Schleswig-Holstein.

### Inclusion criteria

Individuals from the CHOP cohort were included only if they had variants in *DNM1* which were classified as pathogenic or likely pathogenic by a clinical laboratory using the American College of Medical Genetics (ACMG) variant classification criteria or by clinical review from an expert genetic counselor, and for whom medical records were available. Individuals from the literature were included only if they had variants in *DNM1* that were described as pathogenic by the authors of the paper and did not have any alternative or additional identified cause of their neurodevelopmental symptoms. Individuals with variants of uncertain significance in *DNM1* were excluded from analysis.

### Harmonization of clinical features to the HPO

Human Phenotype Ontology terms were manually assigned for all individuals by a genetic counseling student and reviewed by a senior genetic counselor. In addition to “positive” phenotypes (e.g., presence of seizures or autism), “negative” phenotypes (e.g., absence of seizures or absence of abnormal EEG) were also assigned. Negative phenotypes were coded only if absence of the specific phenotypic feature was explicitly documented. For each manually assigned HPO term, automatic reasoning was used to add all applicable higher-level HPO terms up to the root of the HPO ontological tree. This propagation of HPO terms is a well-established method of data harmonization which has been used in several previous studies.^13,14^ We harmonized all HPO data including the data we obtained from literature for other existing datasets to HPO version v2023-06-17.

### Analysis of mutational hotspots

Disease-causing missense variants from individuals in this study were compared to missense variants present with an allele count of at least 1 in the gnomAD v4.1.0 release.^17^ A sliding window approach was used along the primary sequence of the dynamin 1 protein, with window sizes ranging from three to 23 codons. For each window, a positive local likelihood ratio was computed as previously described and the corresponding ACMG evidence weight was assigned.^18^ To each residue, the highest weight across all window sizes was assigned as the final level of evidence.

### Semantic similarity analysis

For each unique HPO term observed in the cohort, we calculated the information content as the -log2 of its frequency within the cohort. The similarity between two individuals was quantified based on the information content of their most informative common ancestor, using the HPO structure as previously described.^13,14^ To assess the significance of overall phenotypic similarity within a given subgroup, we first calculated median similarity score (observed similarity score) among all pairs of individuals in the subgroup. We then created an empirical null distribution by repeatedly selecting random sets of individuals from the overall cohort, matched in size to the subgroup. A total of 100,000 permutations were performed, generating a distribution of median similarity score, resulting in an exact p-value.

## Results

### The genetic landscape of *DNM1* is characterized by recurrent variants

Of the 95 individuals included in the cohort, 90 individuals had heterozygous pathogenic missense variants. There were 45 unique nucleotide variants identified, corresponding to 44 unique amino acid changes. We observed 27 variants in the GTPase domain, 14 variants in the Middle domain, 1 variant in the GED (GTPase effector domain) domain, and 1 variant in the PH (pleckstrin-homology) domain (**Fig. 1A**). No pathogenic variants were observed in the PRD (proline-rich domain). The two most frequently observed variants were p.R237W, which was seen in 15 individuals, and p.I398_R399insCR, which was seen in 14 individuals (**Fig. 1A**). Overall, the two most frequent recurrent variants made up 32.2% of the cohort of individuals with heterozygous pathogenic variants. In addition, we observed 5 individuals that were homozygous for nonsense variants in *DNM1*. Each of the 5 homozygous individuals had unique nonsense variants. Four of the five homozygous individuals came from families with documented consanguinity.

**Figure 1.**
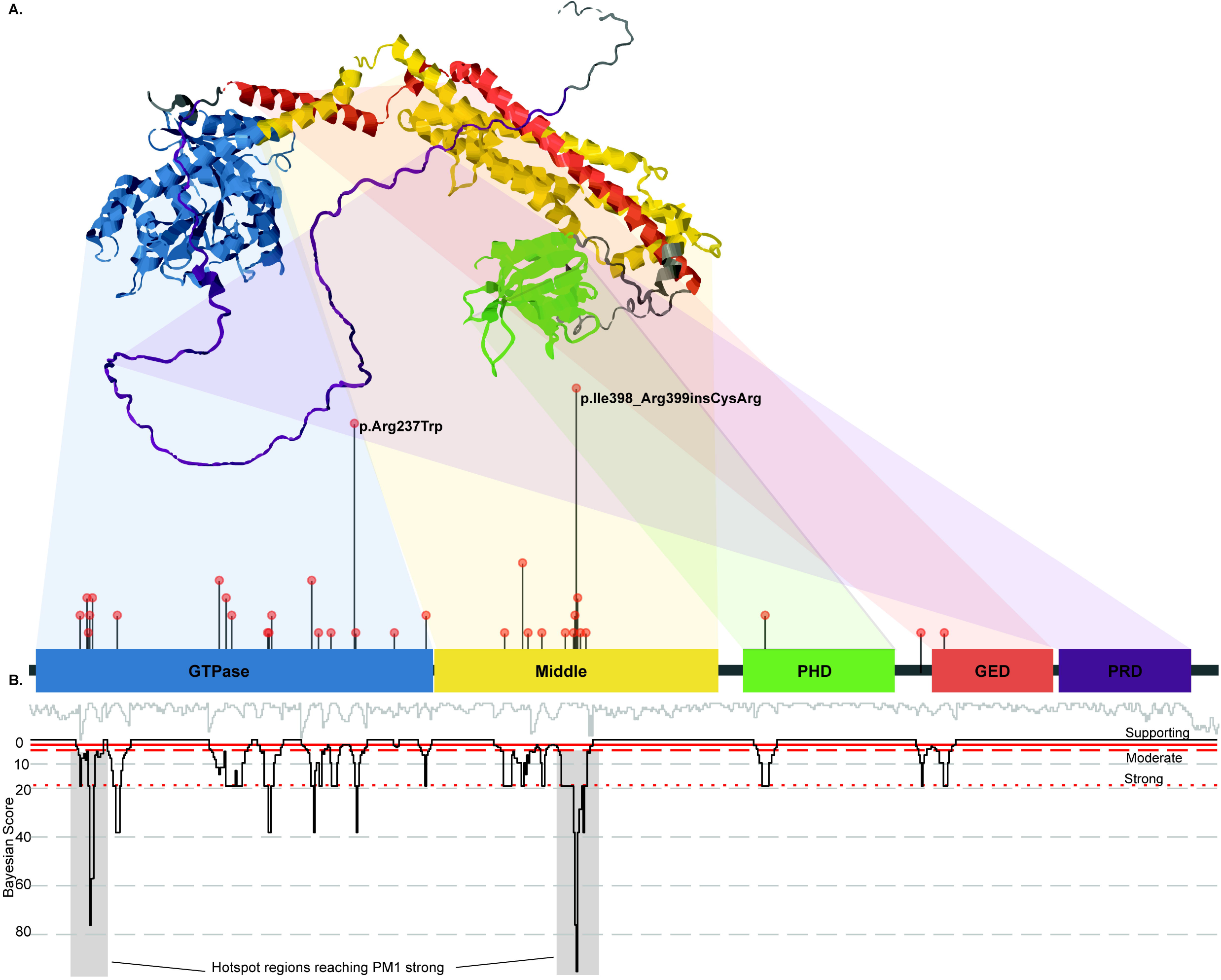
Pathogenic Variants in *DNM1*. **(A)** Distribution of pathogenic missense variants in *DNM1* by genomic position and protein domain, highlighting the two most common recurrent variants. **(B)** Bayesian analysis was used to compare pathogenic and population missense variants to define mutational hotspots within the gene for which the positive local likelihood ratio of pathogenicity is high enough to use as ‘Strong’, ‘Moderate’, and ‘Supporting’ evidence within the ClinGen framework for variant classification.

### ACMG rule-compatible Bayesian analysis defines mutational hotspots

Most pathogenic missense variants in *DNM1* occur in the GTPase and Middle domains, especially at the N-terminus of the GTPase domain and at the start of exon 10a. As part of the ACMG criteria for the classification of variants, points in favor of pathogenicity can be applied to variants which occur in a ‘mutational hotspot’ of the gene. Given the clustering of disease-causing variants in *DNM1* in specific regions of the gene, we assessed whether variant distribution could be mapped to quantitatively define these mutational hotspots, which would be a valuable resource in variant interpretation for *DNM1*-related disorder. Hotspots could be further delineated into ‘supporting’, ‘moderate’, ‘strong’ criterion to be applied in favor of pathogenicity depending on the local likelihood ratio. Bayesian analysis was used to compare 44 disease-causing missense variants to 837 unique population variants from the Genome Aggregation Database (gnomAD v4.1) to calculate the positive local likelihood ratio at each residue of the gene product (**Fig. 1B**). Regions defined by 81 amino acid positions had local positive likelihood ratios greater than 18.71, which represents a mutational hotspot achieving the level of ‘Strong’ evidence for pathogenicity per modified ACMG criteria^18,19^ (**Fig. 1B**). Analogously, a ‘Moderate’ level of evidence per modified ACMG criteria, with positive local likelihood ratio exceeding 4.35, was found for 109 amino acid positions and a ‘Supporting’ level of evidence, with a positive local likelihood ratio exceeding 2.07, was found for 77 amino acid positions^19^ (**Fig. 1B**). A complete list of local likelihood ratios for each amino acid position is available in Supplemental Table 1.

### Standardized clinical terminology captures the clinical spectrum in *DNM1* disorders

We used the Human Phenotype Ontology (HPO) to map the clinical features of *DNM1*-related disorder for quantitative analysis. In total, 1229 HPO terms were assigned across 95 individuals. Harmonization of clinical concepts by adding higher-level clinical terms resulted in a total of 4638 HPO terms. The number of HPO terms per individual ranged from 5 to 115 with a median of 51 terms per individual. The most common terms in the cohort were largely consistent with the phenotype previously reported in the medical literature and reflected a phenotype broadly characterized by epilepsy, hypotonia, movement disorders, and developmental delays (**Fig. 2**). The clinical terms ‘Seizure’, ‘Motor Seizure’, and ‘Epileptic Spasm’ were the most commonly assigned seizure-related terms, which highlights the prevalence and severity of the epilepsy phenotype associated with *DNM1* (**Fig. 2A**). The most common terms related to movement disorders were ‘Hypotonia’ and ‘Abnormality of Tone’, however terms for ‘Involuntary Movements’ and ‘Dystonia’ were also frequently observed (**Fig. 2C**).

**Figure 2.**
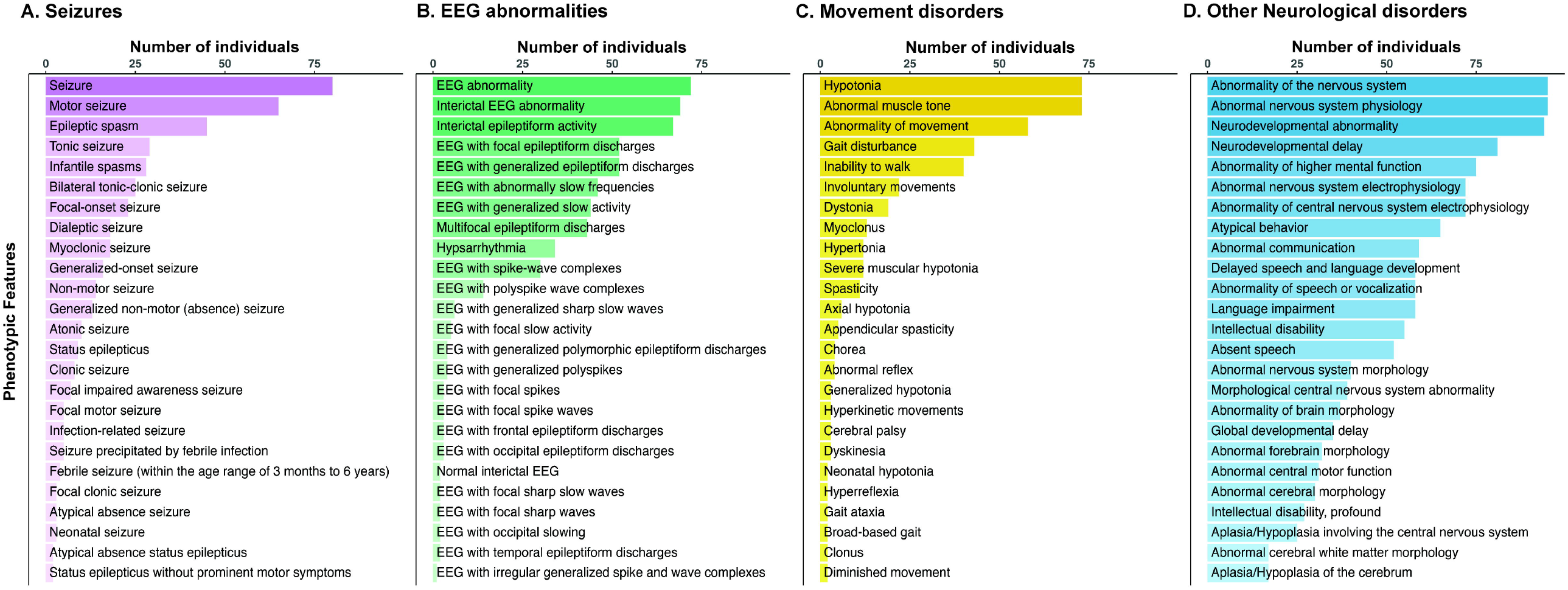
Most frequent HPO phenotype terms in the *DNM1* cohort. **(A)** The Human Phenotype Ontology annotations most frequently assigned in the *DNM1* cohort related to seizure phenotypes **(A)**, EEG abnormalities **(B)**, movement disorders **(C)**, and other neurological symptoms **(D)**. The frequency of terms within this cohort represents a minimum fraction of individuals affected, rather than the overall penetrance of that feature within the cohort, due to the paucity of clinical information available for some included individuals.

### *DNM1*-related disorder is more clinically homogeneous than other genetic epilepsies

Clinical descriptions of rare disorders are typically based on small numbers of individuals and thus comparison to other conditions is a challenge. However, the harmonized framework of the Human Phenotype Ontology allows direct comparison of *DNM1* to other genes for which HPO data sets are available, including *SCN2A, SCN8A, STXBP1*, and *SYNGAP1* (**Fig. 3**). The *SCN2A* cohort included 417 total individuals, 12,787 total propagated HPO terms, and a median of 28 terms per individual.^13^ The *SCN8A* cohort included 359 total individuals and 11,767 total propagated HPO terms.^20^ The *STXBP1* cohort included 534 total individuals, 23,373 total propagated HPO terms, and a median of 41 terms per individual.^14^ And, the *SYNGAP1* cohort included 197 total individuals, 5,796 total propagated HPO terms, and a median of 23 terms per individual.^14^

**Figure 3.**
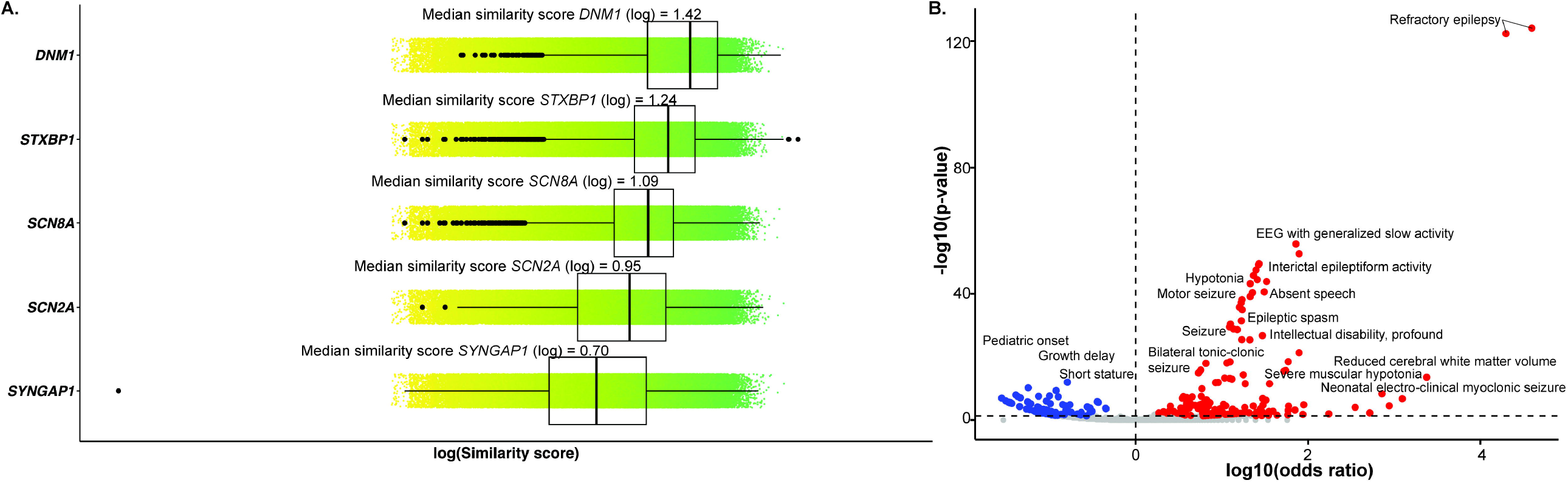
Comparison of *DNM1* to other genetic developmental and epileptic encephalopathies. **(A)** *DNM1*-related disorder has a higher median similarity score than *STXBP1, SCN8A, SCN2A*, or *SYNGAP1*-related disorders, indicating that individuals with *DNM1*-related disorder are more phenotypically similar to one another than are individuals with these other genetic epilepsies. **(B)** Individuals with *DNM1-*related disorder were compared to individuals with *STXBP1, SCN8A, SCN2A*, and *SYNGAP1*-related disorders. Phenotypic terms shown in red with a –log10 (p-value) and a log10 (odds ratio) greater than 0 are enriched in the *DNM1* cohort compared with the other disease cohorts. The top terms associated with DNM1-related disorder were ‘Refractory Drug Response’ (HP:0020174) and its parent term ‘Abnormal Drug Response’ (HP:0020169), which are collectively labeled ‘Refractory epilepsy’ as these terms were applied only to individuals in the cohort with a history of seizures that were refractory to treatment.

Because terms within the Human Phenotype Ontology have defined relationships to one another, and therefore defined distance from one another within a set framework, semantic similarity analysis can be used to capture the degree of similarity or the degree of difference between groups. We utilized semantic similarity analysis to calculate similarity scores across the *DNM1, SCN2A, SCN8A, STXBP1*, and *SYNGAP1* cohorts (**Fig. 3A**). Individuals within the cohort for *DNM1* were significantly similar to one another, as were individuals in the *SCN2A, SCN8A*, and *STXBP1* cohort. In contrast, individuals within the cohort for *SYNGAP1* were not significantly similar to one another. We quantified clinical relatedness within a cohort through median similarity, a measure that captures the degree of overlap in phenotypic terms. The *DNM1* cohort had a median similarity score of 26.5 (p-val = 1.48 × 10^-323^), which was higher than the scores for *STXBP1* (17.7; p-val = 1.17 × 10^-321^), SCN8A (12.5; p-val = 8.79 × 10^-171^), *SCN2A* (8.9; p-val = 3.69 × 10^-64^), or *SYNGAP1* (4.9; p-val =0.107). Phenotypic features which differentiated *DNM1* from the other disorders included EEG with generalized slow activity (HP:0010845; p-val =1.74 × 10^-56^; OR = 72 [95% CI = 46-113]), Hypotonia (HP:0001252; p-val = 5.83 × 10^-44^; OR = 21 [95% CI = 13-51]), and Motor seizure (HP:0020219; p-val = 6.38 × 10^-39^; OR = 17 [95% CI = 10-27]; **Fig. 3B**). The terms Refractory Drug Response (HP:0020174; p-val = 9.23 × 10^-120^; OR = 400 [95% CI = 400-1000]) and its parent term, Abnormal Drug Response (HP:0020169; p-val = 9.23 × 10^-120^; OR = 400 [95% CI = 400-1000]), were also significantly more frequently observed in the *DNM1* cohort. These terms were assigned only to individuals with a documented history of seizures which were refractory to treatment and so can be collectively interpreted as ‘Refractory Epilepsy’ (**Fig. 3B**). In summary, individuals with *DNM1*-related disorder have stronger phenotypic similarity to one another than do individuals with *SCN2A, SCN8A, STXBP1*, or *SYNGAP1*-related disorders.

### Individuals with the most recurrent *DNM1* variants have similar clinical presentations

With the goal of identifying clinically meaningful genotype-phenotype correlations, semantic similarity analysis was performed within the *DNM1* cohort for the two most common recurrent variants: p.R237W (n=15), which falls within the GTPase domain, and p.I398_R399insCR (n=14), which falls in exon 10a in the Middle domain **(Fig. 4A)**. Both recurrent variants had distinct, significantly similar phenotypes, with specific associated features for each genotype. Again, we quantified similarity of phenotypes on a group level through the median similarity score. The p.R237W variant had a median similarity score of 13.67 (p-val = 2.06 × 10^-3^), and the p.I398_R399insCR variant had a median similarity score of 13.32 (p-val = 1.62 × 10^-3^) **(Fig. 4)**. Clinical features associated with the recurrent p.R237W variant included bilateral tonic-clonic seizures (HP:0002069; p-val= 2.82 × 10^-3^; OR = 5.85 [95% CI = 1.6-23.2]), Infantile spasms (HP:0012469; p-val= 1.06 × 10^-2^; OR = 4.7 [95% CI = 1.3-18.4]), and dystonia (HP:0001332; p-val= 1.03 × 10^-2^; OR = 4.8 [95% CI = 1.24-18.8]) **(Fig. 5)**. Clinical features associated with p.I398_R399insCR included severe muscular hypotonia (HP:0006829), profound global developmental delay (HP:0012736; p-val= 1.77 × 10^-9^; OR =124.4 [95% CI = 14.4-5,865]), and cerebral visual impairment (HP:0100704; p-val = 2.43 × 10^-4^; OR = 9.5 [95% CI = 2.5-38.5]) **(Fig. 5)**.

**Figure 4.**
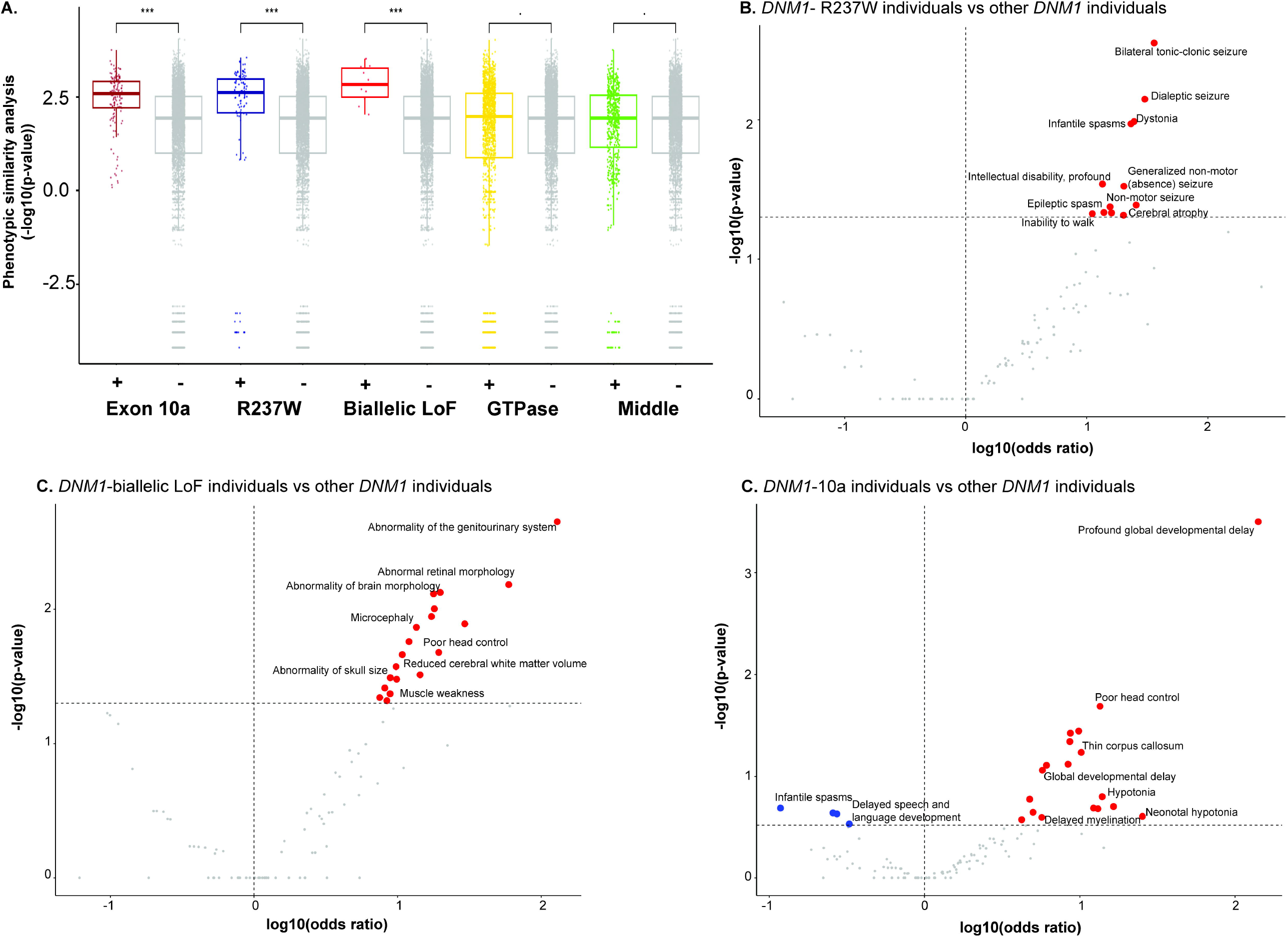
Genotypic subgroups within DNM1-related disorder. **(A)** Semantic similarity analysis for genotypic sub-groups within *DNM1*-related disorder demonstrates that individuals with variants in exon 10a (including p.I398_R399insCR) or the recurrent p.R237W variant are significantly more similar to individuals who share their genotype than to the broader *DNM1* cohort. Individuals with biallelic loss-of-function variants also represent a significantly similar phenotypic cluster. By contrast, individuals with variants in the GTPase domain and Middle domain are not significantly more similar. Phenotypic terms shown in red with a –log10 (p-value) and a log10 (odds ratio) greater than 0 are enriched in individuals with biallelic loss-of-function variants **(B)**, exon 10 variants including the recurrent p.I398_R399insCR variant **(C)**, and the recurrent p.R237W variant **(D)**, respectively.

**Figure 5.**
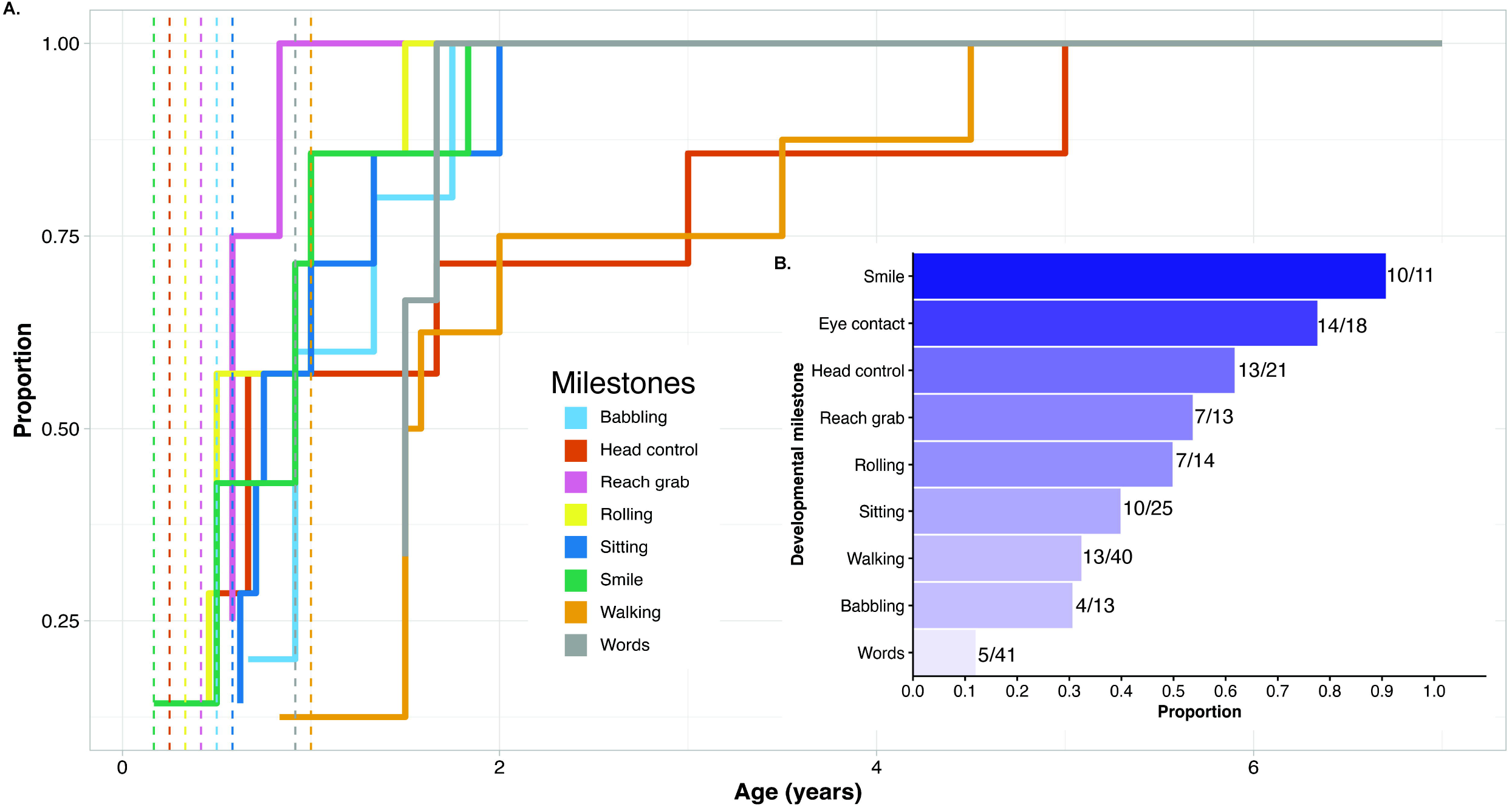
Developmental trajectories in *DNM1-*related disorder. **(A)** Where records were available, the age at which individuals attained developmental milestones was extracted. Curves demonstrate that those individuals who attained each milestone within the DNM1 cohort did so later than would be expected for a typically developing individual. Typical ages of milestone attainment are represented by vertical lines and based on Scharf et al.^49^ **(B)** Proportion of individuals in the cohort meeting each milestone who are above the typical age of attainment for that milestone and for whom explicit records of developmental attainment are available.

By contrast, phenotypes for variants in the GTPase and Middle domains, when taken together, were not significantly similar to one another **(Fig. 5)**. In addition, no significant genotype-phenotype correlations were associated with these domains as a whole.

### Biallelic loss-of-function is clinically similar to dominant variants with some distinct features

The initial cases of *DNM1* reported in the literature identified heterozygous missense variants and suggested that the mechanism of pathogenicity is a dominant-negative effect.^8^ However, subsequent case reports identified a clinically similar phenotype in several individuals with biallelic protein-truncating variants in *DNM1* inherited in trans from unaffected parents who were each heterozygous for the variant, suggesting that biallelic loss-of-function is an additional mechanism of disease associated with *DNM1*.^10-12^ To assess the phenotypic similarity of these groups, we compared the phenotypes of recessively inherited loss-of-function variants to dominantly inherited missense variants. Similarity analysis within the *DNM1* cohort for the individuals with biallelic loss-of-function variants (n=5) showed a median similarity score of 17.08 (p-val =1.07 × 10^-2^) **(Fig. 4)**. This indicates that individuals with biallelic loss-of-function variants represent a distinct cluster within the *DNM1* cohort **(Fig. 4)**. Notable phenotypic terms most associated with biallelic loss-of-function variants were microcephaly (HP:0000252; p-val = 1.75 × 10^-2^; OR = 11.4 [95% CI = 1.1-152.6]), motor delay (HP:0001270; p-val = 1.74 × 10^-2^; OR = 11.4 [95% CI = 1.1-152.6]), and poor head control (HP:0002421; p-val = 2.67 × 10^-2^; OR = 9.4 [95% CI = 0.97-123]), as well as several terms related to abnormal brain morphology **(Fig. 5)**. Abnormality of the genitourinary system also emerged as a significant term, however this is likely an artifact of the phenotyping process, rather than a clinically significant phenotypic difference, since 2 of the 5 homozygous individuals were assigned the term for Urinary incontinence (HP:0000020). Overall, individuals with biallelic loss-of-function variants had a similar clinical presentation to the overall DNM1 cohort, with a phenotype characterized by intractable epilepsy and severe developmental delays. However, the top terms associated with biallelic-loss-of function suggest that individuals with this genetic mechanism are more likely to present with microcephaly and significant brain MRI findings.

### Developmental outcomes in *DNM1* are characterized by global delays without regression

Previous work has demonstrated that *DNM1-*related disorder is associated with severe-profound developmental delays and intellectual disability.^8^ The proportion of individuals attaining particular developmental milestones was extracted from information available in either the medical records or the literature **(Fig. 6A)**. Individuals were included if they were older than the age at which that milestone would be achieved by a typically developing child, and only if information about that milestone was specifically recorded **(Fig. 6B)**. A majority of individuals achieved a responsive smile, head control, babbling, and rolling. All other milestones were attained by less than 50% of the cohort. 40% of individuals were able to sit independently, 32.5% were able to ambulate independently, and 12% had some degree of verbal communication. Developmental regression did not emerge as a significant feature of *DNM1*-related disorder, the HPO term for developmental regression was assigned only for eight individuals. The age at which individuals attained developmental milestones was also extracted where available **(Fig. 6A)**. For those individuals who were ambulatory, the median age of walking was 1.54 years (range 10 months t0 4.5 years). For individuals who had verbal speech, two individuals had first words at 18 months and one individual at 20 months. Information about the age of first words was otherwise not available.

## Discussion

Here, we used a data analytics approach to circumvent the challenges innate to small sample sizes in rare disease research. A combined data set of 95 participants was drawn from a heterogenous set of sources with highly variable degrees of phenotype information, and the HPO framework was utilized to create a harmonized data set large enough to construct a more nuanced picture of the *DNM1* phenotype. We analyzed the distribution of pathogenic and benign variants across the gene transcript to define mutational hotspots in a practical manner that can be used to guide future variant classification. The relational nature of the HPO made it possible to analyze the overall phenotypic landscape in comparison to other genetic epilepsies: *SCN2A, SCN8A, STXBP1*, and *SYNGAP1*. We also investigate genotype-phenotype correlation for recurrent missense variants and biallelic loss-of-function variants to provide clinical insights.

### Classification of missense variants in *DNM1*

*DNM1*-related disorder poses a particular challenge in distinguishing pathogenic variants from benign variants. Consistent with its dominant-negative mechanism, the majority of pathogenic variants in *DNM1* are missense variants. Loss-of-function is not a known disease mechanism, except in the biallelic state. Accordingly, it is critical to differentiate between pathogenic and benign missense variation. Variants of uncertain significance can be deeply frustrating for families and add to the complexity of clinical decision-making. We used an ACMG/AMP compliant Bayesian analysis to map the pathogenic and benign missense variants in *DNM1* to statistically define the mutational hotspots that correlate with ‘Strong’, ‘Moderate’, and ‘Supporting’ evidence of pathogenicity as laid out by the ACMG guidelines for variant classification. These results and the identified variant hotspot regions will aid in the classification of future novel missense variants in *DNM1*.

### Implications for clinical management of individuals diagnosed with *DNM1*-related disorder

The aim of this study was to clarify the core phenotype of *DNM1*-related disorder in order to take initial steps towards clinical trial readiness in this condition and allow for more concrete clinical guidance to families. The core clinical findings are hypotonia (73 individuals), seizures (80 individuals), and severe to profound intellectual disability (15 and 27 individuals respectively). The epilepsy associated with *DNM1* is characterized multiple seizure types, but frequently presented with infantile spasms. The median age of onset for seizures was 5 months but ranged from day-of-life 5 to 16 years. While seizures were a core phenotype feature, there were 12 individuals (12.6%) who were seizure-free at last evaluation. Most individuals had seizures that were refractory to multiple anti-seizure medications with 54% of the cohort explicitly documenting refractory seizures, and no specific medications have been shown to be more effective in this population at this time.

Individuals with *DNM1*-related disorder generally have severe to profound intellectual disability. Of the developmental milestones assessed, only responsive smile, head control, babbling, and rolling were achieved by a majority of participants for whom that milestone was age-appropriate and information was available. Of the individuals older than 1 year of age, 32% were able to ambulate independently and 12% had some degree of verbal communication. Participants in this study ranged in age from 10 months to 24 years, and there is no clear evidence that *DNM1*-related disorder is a progressive condition.

An exception to the phenotype described above is individuals with variants in exon 10b of *DNM1*, which is less expressed in brain tissue.^9^ One individual with a pathogenic variant in exon 10b of *DNM1* has been observed with a less severe phenotype that included mild global developmental delay without clinical seizures by age 4.^9^ This notable case has been demonstrated, but it is possible that other novel missense variants identified in the future might be associated with more mild disease. While we demonstrate the homogeneity of the *DNM1* phenotype, the exon 10b case demonstrates that there are exceptions.

For individuals diagnosed with pathogenic variants in exon 10a of *DNM1*, including the recurrent p.I398_R399insCR variant, more guidance about clinical features can be tailored to this specific genotype. While developmental delay was a common feature across genotypes, profound intellectual disability was significantly more common in individuals with exon 10a variants, as well as severe hypotonia and poor head control. There were also several clinical features associated with the recurrent p.R237W variant. Individuals with this variant had a higher prevalence of infantile spasms and were more likely to present with bilateral tonic-clonic seizures. Individuals with the p.R237W variant were also more likely to have movement disorders, specifically dystonia. For individuals diagnosed with biallelic loss-of-function variants in *DNM1*, the phenotype is broadly similar to the phenotype of autosomal dominant *DNM1*-related disorder with regards to epilepsy and development. However, individuals with this genotype had a higher prevalence of microcephaly and brain MRI abnormalities.

This work draws upon a large number of sources of heterogeneous clinical information. While this approach confers many advantages, it also has some limitations. The degree of phenotypic information available for each included individual varied widely. The use of HPO terms to standardize the data set helped to minimize the impact of reporting or recruitment bias within individual papers. However, due to the fact that some of the individuals included in the cohort had a very limited phenotype information available, percentages of traits within this cohort represent a minimum percentage affected, rather than the overall penetrance of that feature within the cohort. We also acknowledge that the cohort of patients is likely enriched for individuals with the recurrent p.I398_R399insCR variant due to a previous study published at the institution which specifically recruited individuals with variants in exons 10a and 10b. For this reason, we do not draw a broader conclusion about the prevalence of the p.I398_R399insCR variant among individuals with *DNM1*-related disorder.

### *DNM1* may be a promising target for the development of targeted therapies

DNM1-related disorder has several characteristics which may aid in the development of novel, targeted therapies. The mechanism of disease is dominant-negative, which makes antisense-oligonucleotide-based therapies a potentially effective option for individuals with heterozygous pathogenic variants. A relatively high proportion of patients have one of two recurrent variants, so it would be possible to develop genotype-specific treatments that would be usable for a large proportion of affected individuals. The homogeneity of the core phenotype may also make it possible for clinical trials to utilize a natural history arm rather than relying on a placebo control arm. Clear characterization of the phenotype and natural history of *DNM1*-related disorder will be necessary for clinical trial readiness for *DNM1*.

In summary, our study provides wider insights about the way that *DNM1* fits into the larger landscape of genetic epilepsies. We characterize mutational hotspots and phenotypic features associated with the most recurrent variants. In addition, it will also provide granular clinical insights that might be helpful to individual patients and families. A clearer understanding of the phenotype of this rare disorder might eventually provide the foundation to search for effective interventions.

## Data Availability

The datasets and code generated during this study are available at https://github.com/helbig-lab/dnm1-hpo.

## Abbreviations

ACMG: American College of Medical Genetics and Genomics
ASM: anti-seizure medications
CHOP: Children’s Hospital of Philadelphia
DEE: developmental and epileptic encephalopathy
EOEE: early onset epileptic encephalopathy
HPO: Human Phenotype Ontology
NDD: neurodevelopmental disorders

## Data and Code Availability

The datasets and code generated during this study are available at https://github.com/helbig-lab/dnm1-hpo. The HPO data set is included as Supplemental Table 2.

## Acknowledgements

We would like to thank patient advocates from the DNM1 Epilepsy foundation for their support and contribution to patient recruitment. We additionally thank Annalaura Torella, Vincenzo Nigro, Lino Nobili, and Federico Zara for their help in participant recruitment. The authors also thank the patients and families who selflessly contributed their time and medical information to this study.

## Author Contributions

A.H. contributed to investigation, data curation, and writing (original draft). S.G., M.X., S.P., and J.H.M. contributed to methodology, formal analysis, data curation, visualization, and writing (review and editing). J.L.M contributed to methodology, resources, and writing (review and editing). K.T. and R.L. contributed to investigation and data curation. M.P., A.B., H.M., A.P., A.M., G.W., K.O., A.L.B., and M.S. contributed to resources and writing (review and editing). S.R. contributed to conceptualization, methodology, resources, supervision, and writing (review and editing). I.H. contributed to conceptualization, methodology, supervision, funding acquisition, and writing (review and editing).

## Ethics Declaration

This study was reviewed by the Children’s Hospital of Philadelphia Institutional Review Board (IRB) as part of the Epilepsy Genetics Research Project (EGRP) 15-12226. All institutions involved in participant recruitment received local IRB approval. Informed consent was obtained from all participants. Data provided to the Children’s Hospital of Philadelphia from collaborators was de-identified.

